# Recommendations for clinical interpretation of variants found in non-coding regions of the genome

**DOI:** 10.1101/2021.12.28.21267792

**Authors:** Jamie M Ellingford, Joo Wook Ahn, Richard D Bagnall, Diana Baralle, Stephanie Barton, Chris Campbell, Kate Downes, Sian Ellard, Celia Duff-Farrier, David R FitzPatrick, Jodie Ingles, Neesha Krishnan, Jenny Lord, Hilary C Martin, William G Newman, Anne O’Donnell-Luria, Simon C Ramsden, Heidi L Rehm, Ebony Richardson, Moriel Singer-Berk, Jenny C Taylor, Maggie Williams, Jordan C Wood, Caroline F Wright, Steven M Harrison, Nicola Whiffin

## Abstract

**Purpose:** The majority of clinical genetic testing focuses almost exclusively on regions of the genome that directly encode proteins. The important role of variants in non-coding regions in penetrant disease is, however, increasingly being demonstrated, and the use of whole genome sequencing in clinical diagnostic settings is rising across a large range of genetic disorders. Despite this, there is no existing guidance on how current guidelines designed primarily for variants in protein-coding regions should be adapted for variants identified in other genomic contexts.

**Methods:** We convened a panel of clinical and research scientists with wide-ranging expertise in clinical variant interpretation, with specific experience in variants within non-coding regions. This panel discussed and refined an initial draft of the guidelines which were then extensively tested and reviewed by external groups.

**Results:** We discuss considerations specifically for variants in non-coding regions of the genome. We outline how to define candidate regulatory elements, highlight examples of mechanisms through which non-coding region variants can lead to penetrant monogenic disease, and outline how existing guidelines can be adapted for these variants.

**Conclusion:** These recommendations aim to increase the number and range of non-coding region variants that can be clinically interpreted, which, together with a compatible phenotype, can lead to new diagnoses and catalyse the discovery of novel disease mechanisms.

## INTRODUCTION

Genomic sequencing is commonplace in the diagnosis of disorders with suspected genetic cause. Traditionally, sequencing and analysis has focussed primarily on variants that fall within regions of the genome that code directly for protein, or that are within canonical splice-sites of genes with a confirmed role in disease. With these approaches, however, many rare disease cases remain genetically unexplained^1,2^.

Increasingly, whole genome sequencing (WGS) is being performed on individuals in which a genetic cause is not identified through gene panel or exome sequencing. For some disease subsets and in specific healthcare settings, WGS has become a first line diagnostic test (for example for selected rare developmental disorders in the UK National Health Service)^3^. WGS has been shown to have the potential to increase diagnostic yield^2,4–7^, and includes detection of variants in a wide-range of regulatory regions (Box 1) as well as variants in genes encoding non-coding RNAs (e.g. micro RNAs (miRNA), small nuclear RNAs (snRNA), and long non-coding RNAs). Analysis of WGS data, however, often excludes variants that fall in non-coding regions of the genome or classifies them as variants of uncertain significance (VUS), primarily due to difficulties in predicting or determining their impact. Whilst the triplet amino acid code allows us to predict the effect of variants within protein-coding regions with reasonable accuracy, the absence of a regulatory equivalent means that the impacts of non-coding region variants are usually much harder to predict. This is further confounded by these variants often having gene-specific effects. For example, binding sites for the zinc finger protein CCCTC-binding factor (CTCF) are enriched at topologically associated domain (TAD) boundaries and have been suggested as a mechanism to ensure appropriate genome regulation and chromatin structure. However, it is unclear why in some instances disrupting CTCF binding sites significantly impacts gene expression^8^, and in others it does not^9,10^. Such differences may be dependent on the surrounding genomic context and the temporal/spatial activity of other cis-regulatory elements (CREs)^11^.

An important role for a range of non-coding variants in rare disease is increasingly being demonstrated^12–14^. For example, variants in upstream non-coding regions that cause loss-of-function of *MEF2C* comprise almost one quarter of all likely diagnoses impacting *MEF2C* in the Deciphering Developmental Disorders (DDD) dataset^15^, and RNA-sequencing can be used to identify likely disease-causing splicing variants in 35% of previously undiagnosed rare muscle disease probands, many in deep intronic regions^16^. A number disease-causing variants for X-linked Charcot-Marie-Tooth disease in *GJB1*^*17*^ and ABCA4-associated disease^18,19^ are also known to be in non-coding regions.

There are a multitude of documented mechanisms through which non-coding region variants which disrupt non-coding elements have been demonstrated to cause severe disease (Table 1). These include acting through affecting splicing^16,24,25^, transcription^26,27^, translation^15^, RNA processing and stability^12,28^, and chromatin interactions^29^. Detecting and classifying these variants accurately for a likely disease-causing role is important to increase diagnostic yield and enable a robust genetic diagnosis for more individuals.

**Table 1:** Categories of small variants in non-coding regions previously implicated in penetrant Mendelian disease. This is not intended as an exhaustive list. Reference DOIs: 1. 10.1182/blood-2018-07-863951. 2. 10.1002/humu.23212. 3. 10.1016/j.ajhg.2018.07.002. 4. 10.1016/j.ebiom.2016.04.005. 5. 10.1038/s41467-019-10717-9. 6. 10.1074/jbc.M005199200. 7. 10.1038/s41431-020-0676-y. 8. 10.1002/ajmg.a.36703. 9. 10.1016/j.ajhg.2021.04.025. 10. 10.1172/JCI119555. 11. 10.2337/db07-1657. 12. 10.3390/genes11101180. 13. 10.1016/j.ajhg.2018.10.023. 14. 10.1136/jmedgenet-2020-107626. 15. 10.1016/j.ajhg.2019.07.013. 16. 10.1182/blood-2018-03-838235. 17. 10.1136/jmedgenet-2018-105836. 18. 10.1086/505361. 19. 10.1038/s10038-018-0502-3. 20. 10.1038/ng.230. 21. 10.1038/ng.2826. 22. 10.1038/ng.329. 23. 10.1038/s41467-021-23980-6. 24. 10.1073/pnas.1401464112. 25. 10.1002/ana.24826. 26. 10.1038/ncomms9718. 27. 10.1038/ng.3661. 28. 10.1016/j.cell.2015.04.004.

| Region | Mechanism | Example (variant/gene) | Example (disease) | ClinVar Var ID | Reference | VEP categories | In silico tools to predict effect |
| --- | --- | --- | --- | --- | --- | --- | --- |
| Promoter | Altering transcription factor binding | GATA1:-113A>G | Hereditary persistence of fetal hemoglobin |  | 1 | upstream gene variant / regulatory region / TF binding site | TF binding site disruption prediction tools e.g. motifbreakR / SEMpl / QBiC-Pred |
| Promoter | Altering transcription | CHM c.-98C>A, c.-98C>T | Choroideremia |  | 2 |  |  |
| Promoter/5'UTR | Altering methylation patterns | BRCA1:c.-107A>T | Breast and Ovarian Cancer |  | 3 | upstream gene variant / 5 prime UTR variant |  |
| 5'UTR | Creating upstream start site (uAUG) | NF1:c.-272G>A | Neurofibromatosis type 1 | 1013130 | 4 | 5 prime UTR variant | e.g. UTRannotator |
| 5'UTR | Perturbing upstream open reading frames | NF2:-66-65insT | Neurofibromatosis type 2 |  | 5 |  | e.g. UTRannotator |
| 5'UTR | Disrupting internal ribosome entry sites (IRES) | GJB1:c.-103C>T | Charcot-Marie-Tooth disease | 217166 | 6 |  | e.g. IRESpy / IRESfinder / IRESite |
| 5'UTR | Disrupting splicing | SHOX:c.-19G>A | SHOX haploinsufficiency | 933226 | 7 |  | Splicing prediction tools e.g. SpliceAI |
| 5'UTR | Altering Kozak consensus of start site | GATA4:c.-6G>C | Atrial septal defect |  | 8 |  | e.g. utR.annotation |
| 5'UTR | N-terminal transcript elongation | MEF2C:c.-8C>T | Developmental disorder |  | 9 |  | e.g. UTRannotator |
| Intron | Disrupting canonical splice sites | MYBPC3:c.3490+1G>A | Hypertrophic cardiomyopathy | 42715 | 10 | splice donor variant / splice acceptor variant / splice region variant | Splicing prediction tools e.g. SpliceAI |
| Intron | Disrupting splicing branch point | HNF4A:c.264-21A>G | Maturity-onset diabetes of the young |  | 11 | intron variant | Splicing prediction tools e.g. SpliceAI |
| Intron | Pseudo-exon activation | DMD: c.7310-19A>G | Muscular dystrophy |  | 12 |  | Splicing prediction tools e.g. SpliceAI |
| Intron | Poison-exon inclusion | SCN1A:c.4002+2165C>T | Dravet Syndrome |  | 13 |  | Splicing prediction tools e.g. SpliceAI |
| Intron | Branchpoint mutation | BBS1:c.592-21A>T | Retinitis pigmentosa |  | 14 |  | Splicing prediction tools e.g. SpliceAI |
| Intron | Indels & spacing of splicing motifs | DOK7:c.54+8_54+17del |  |  | 15 |  | Splicing prediction tools e.g. SpliceAI |
| Intron | Cryptic exon | VHL: c.3401770T.C | Erythrocytosis |  | 16 |  |  |
| 3'UTR | Disrupting polyA signal motif | NAA10:c.*43A>G | Microphthalmia | 617463 | 17 | 3 prime UTR variant | PolyA signal motif prediction tools e.g. Omni-PolyA |
| 3'UTR | Disrupting miRNA interactions | REEP1 | Hereditary Spastic Paraplegia |  | 18 |  | miRNA binding site prediction e.g. miRTarBase |
| 3'UTR | Disrupting splicing | LHFPL5:c.*16+1G>A | Hearing impairment |  | 19 |  | Splicing prediction tools e.g. SpliceAI |
| CRE | Altering transcription factor binding | chr7:155754267:C>T; NCBI build 36.3 | Holoprosencephaly |  | 20 | upstream gene variant / downstream gene variant / regulatory region variant / TF binding site variant / intergenic variant / TFBS ablation / TFBS aplication / regulatory region ablation / regulatory region aplication | TF binding site disruption prediction tools e.g. motifbreakR / SEMpl / QBiC-Pred |
| CRE | Abolishing enhancer activity | PTF1A - 6 variants | Isolated pancreatic agenesis |  | 21 |  |  |
| CRE | Disrupting enhancer activity | SOX9 - deletion (chr17:67,628,756–67,634,155) | Pierre Robin sequence |  | 22 |  |  |
| Intergenic | Creating new regulatory element | chr16:209,709 T>C | α-thalassaemia | | 23 | intergenic variant / upstream gene variant / downstream gene variant | |
| miRNA | Disrupting seed region | miR-204:n.37C>T | Retinal dystrophy |  | 24 | non coding transcript exon variant / non coding transcript variant |  |
| snRNA | Altering structure | RNU12 | Cerebellar ataxia |  | 25 | non coding transcript exon variant / non coding transcript variant | Splicing prediction tools e.g. SpliceAI |
| snRNA | Abnormal splicing, accumulation of minor intron retained transcripts | RNU4ATAC | Roifman Syndrome |  | 26 |  |  |
| snRNA | Affecting expression, processing and protein binding | SNORD118 | Cerebral microangiopathy leukoencephalopathy |  | 27 |  |  |
| TAD boundary | Disrupting chromatin looping leading to enhancer loss or adoption | WNT6/IHH/EPHA4/PAX3 locus | Limb phenotypes |  | 28 | intergenic variant |  |

The American College of Medical Genetics and Genomics and Association for Molecular Pathology (ACMG/AMP) released a set of guidelines in 2015 that have become the global standard for interpreting the pathogenicity of short sequence variants (single nucleotide variants (SNVs) and indels <50bps) identified in individuals with rare disease^30^. These guidelines outline a set of rules that should be assessed for each identified variant. Many of these rules pertain specifically to variants in protein-coding regions and there is no existing guidance on how they should be adapted for variants found in other genomic contexts. Here, we provide guidance on how to apply these standards to variants identified in non-coding regions of the genome. Our recommendations will enable consistent interpretation and reporting of these understudied variant types which will in turn enable us to learn more about the diverse mechanisms through which non-coding region variants can lead to disease.

### Box 1: Regulatory elements controlling gene and protein expression

**Figure 1:**
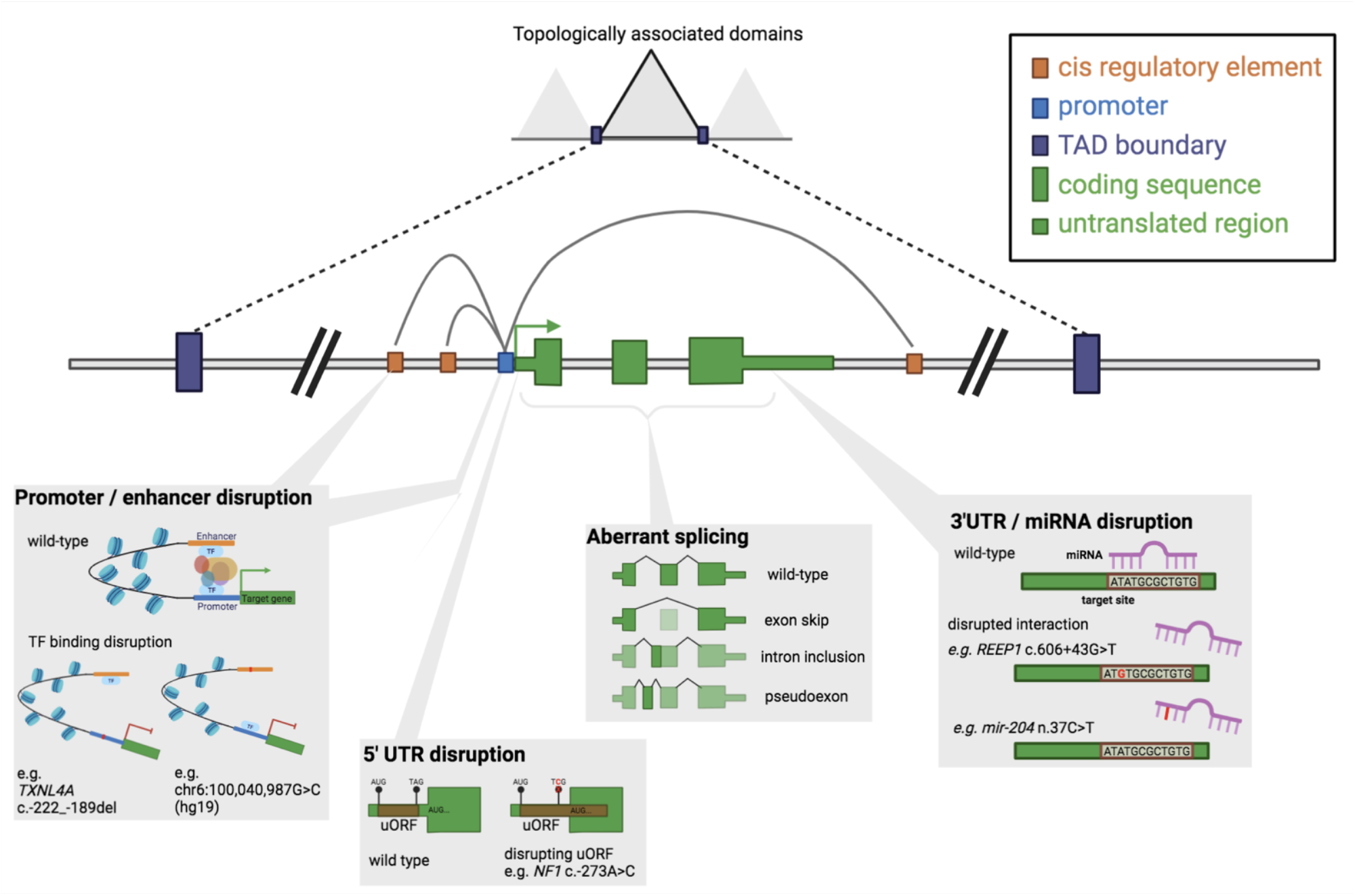
Schematic of regulatory elements within and around a gene and examples of disruptions that can lead to disease. Gene and protein expression are tightly controlled processes mediated by a multitude of regulatory elements (Figure 1). Transcription of a gene into RNA is mediated by a promoter element directly upstream of the gene^20^, along with more distal enhancer and repressor elements (collectively referred to as cis-regulatory elements, or CREs) to which transcription factors bind^21^. CREs may be within their target gene or in other intragenic or intergenic space either 3’ or 5’ of the transcription unit they influence. Within the gene itself, intronic regions contain specific sequences that control their removal through splicing to form the mature RNA (mRNA) transcript, and untranslated regions (UTRs) regulate RNA stability, trafficking, and the rate at which it is translated into protein^22^. Each gene also sits within a wider regulatory context, or topologically associated domain (TAD), flanked by boundary/insulator elements which restrict the action of CREs to within specific TADs^21,23^.

## METHODS

### Process for drafting and refining the recommendations

We convened a panel of clinical and research scientists with wide-ranging expertise in clinical variant interpretation, with specific interests and experience in variants within non-coding regions. The initial document was drafted and circulated before a series of online calls to discuss and refine the guidance. Subsequently, we asked a range of clinical scientists and those actively involved in clinical variant interpretation to test the usability of the guidelines on both a common list of 30 diverse variants (Supplementary Table 1) and in-house identified variants. Feedback from testers was used to further refine the guidance.

At the Association for Clinical Genomic Science (ACGS) meeting in September 2021 we presented an overview of the new guidelines to the UK Clinical Science community. At this meeting we polled opinions on current best practice, the need for specific guidelines for non-coding region variants, and the appetite for training workshops/seminars (Supplementary Table 2). The attendees of the workshop were overwhelmingly in support of this effort with 98% (59/60) agreeing that additional guidance is needed to support interpretation of non-coding region variants.

### Assessing non-coding region variants in ClinVar

To identify non-coding region variants in ClinVar^31^, all variants (n=789,941) from the ClinVar GRCh38 VCF dated 01/31/2021 were annotated with respect to MANE Select v0.93 transcripts. Variants were assigned as falling within the coding sequence (n=597,408), 5’UTR (n=10,820), 3’UTR (n=53,988), intronic regions (n=110,618), or in the 2kb upstream of the transcription start site (annotated as promoter; n=4,348). All remaining variants (n=12,759) were assigned as ‘other’. The majority of these ‘other’ variants were coding sequence variants in genes without designated MANE Select transcripts.

High-confidence pathogenic variants were designated as those with a review status of ‘criteria_provided,_multiple_submitters,_no_conflict s’, ‘reviewed_by_expert_panel’, or ‘practice_guideline’. Pathogenic variants were taken as those with significance of ‘Pathogenic’, ‘Likely_pathogenic’, or ‘Pathogenic/Likely_pathogenic’.

### Identifying in trans non-coding region variants in 100,000 Genomes Project

To determine the frequency of variants observed *in trans* with potentially pathogenic variants, we used the Genomics England (GEL) 100,000 Genomes dataset (version 7). We identified all probands recruited as full trios (i.e. an affected proband and both unaffected parents) without variants classified as either tier 1 or tier 2 in the GEL clinical filtering pipeline (https://research-help.genomicsengland.co.uk/pages/viewpage.action?pageId=38046769). We next identified all remaining probands with a single heterozygous predicted loss-of-function (pLoF) variant in one of 794 genes catalogued as biallelic loss-of-function (LoF) genes in the Developmental Disorders Gene to Phenotype (DDG2P) database^32^ (downloaded on 02/04/2019). Variants were filtered to only those classified as high-confidence by LOFTEE^33^, with allele frequency (AF) <0.5% across the GEL rare disease cohort and/or in gnomAD v2.1.1^33^, and with >25% but <75% of reads containing the variant.

Each DDG2P biallelic LoF gene was annotated with a minimal set of non-coding regulatory regions comprising all intronic regions, the 5’UTR and 3’UTRs, and a core promoter region comprising the first 200 bps directly upstream of the transcription start site. Regions were identified using the MANE Select v0.9 transcript where available, and otherwise the canonical transcript as defined by UCSC. Variants transmitted by the alternative parent to the pLoF variant were identified in the non-coding regions of the same gene and filtered to only those with filtering allele frequency^34^ <0.5% and no observed homozygotes in gnomAD v3.1.

## RESULTS

### Non-coding region variants are under-ascertained in clinical variant databases

Non-coding regions are not regularly captured in clinical sequencing pipelines, where they are most often excluded from capture regions, or removed during bioinformatic processing of the data. Consequently, even variants within non-coding regions known to regulate established disease genes are under-reported. In the ClinVar database^31^ only 1 in 294 (0.34%) high-confidence pathogenic variants are in UTRs or immediately upstream regions (within 2kb; Figure 2). This is despite UTRs having approximately the same genomic footprint as protein-coding regions and important regulatory roles^28^. Whilst this is in part due to the lower likelihood of any single non-coding region variant being pathogenic, it also reflects under-ascertainment.

**Figure 2:**
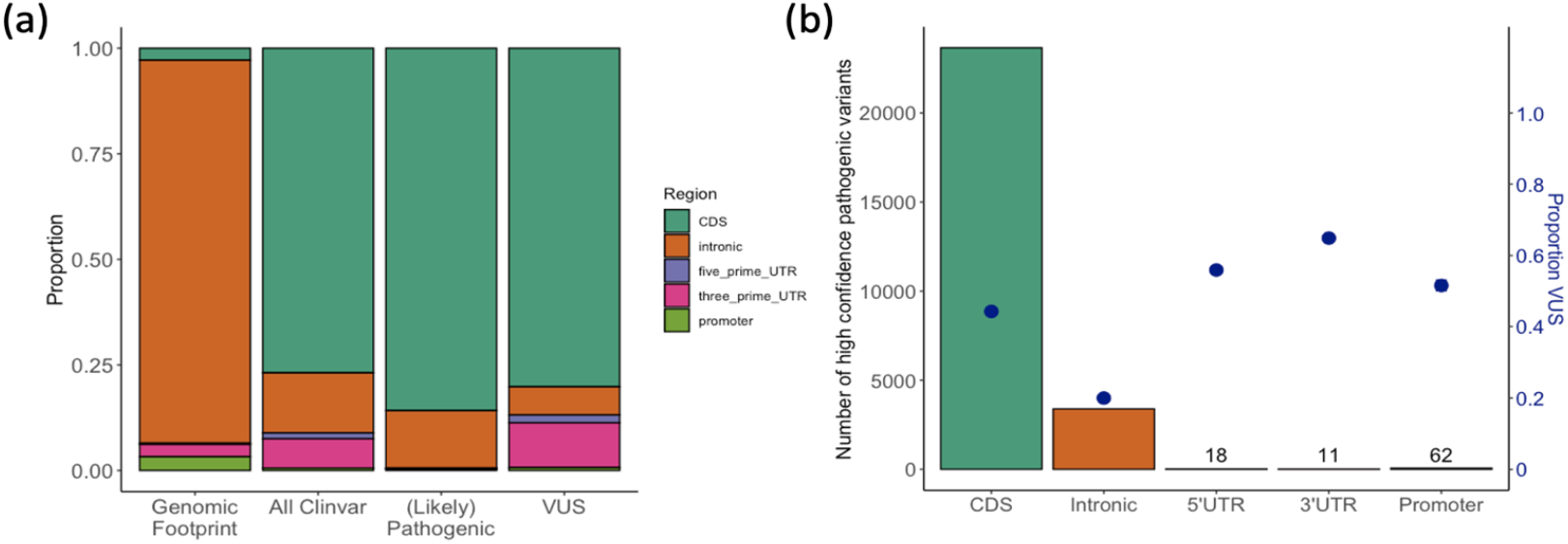
Non-coding region variants are under-ascertained in ClinVar and are more likely to be classified as variants of uncertain significance (VUS) when compared to protein coding variants. (a) The proportion of the genomic footprint of MANE transcripts that fall into each of five region categories and the proportion of variants in ClinVar (all, likely pathogenic or pathogenic, and VUS) within those regions. (b) The number of high confidence pathogenic variants in ClinVar (see methods) that fall into each of the five region categories plotted as bars, with the proportion of variants in each region classified as VUS as blue points.

Regulatory variants are also more likely to be categorised as variants of uncertain significance (VUS), with 63.4% of all UTR variants in ClinVar categorised as VUS, compared to 44.2% of coding sequence variants (Figure 2b), highlighting the need for clearer guidelines for interpretation and strategies for functional validation.

### Defining and filtering candidate non-coding regions

The vast majority of the thousands of non-coding region variants identified in each individual will have very little or no effect. For example, whilst 43% of all assessed common variants (minor allele frequency ≥0.01) have a significant effect on expression of at least one gene in at least one human tissue, 78% of these confer <2-fold changes^35^. To avoid both a huge burden of interpretation and many variants being reported as variants of uncertain significance (VUS), it is important to only clinically interpret variants that (1) fall into regulatory elements that have well-established or functionally validated links to target genes, and (2) those genes have documented associations to the phenotype of interest (i.e. at definitive, strong or moderate level using the ClinGen classification approach^36^, or green for the phenotype of interest in PanelApp^37^). This should be used as a filtering approach rather than evidence towards pathogenicity. We therefore only recommend the use of ACMG/AMP rule PP4 for non-coding region variants where the gene is the only, or one of very few genes associated with a discriminative set of phenotypic features, in accordance with existing guidance^30^. For variants within candidate CREs or non-coding genes without proven gene-disease validity we recommend that they are treated as research variants, and not interpreted or reported, until meaningful functional experiments prove a direct effect on the target gene.

We note that identifying CREs and linking them to specific genes is a very active area of research^21,38^. However, based on current knowledge we recommend that regions of interest within which variants should be interpreted should be defined using the following parameters. Possible sources of data to support these definitions are listed in Table 2.

**Table 2:** Data types used for identification of candidate non-coding regions. *https://doi.org/10.1186/s13059-018-1519-9; ^Tissue specificity and temporal specificity (e.g. specific to a developmental time point) should be considerations for all.

| Evidence | Description | Region(s) | Possible source | Extra considerations^ |
| --- | --- | --- | --- | --- |
| ATAC-seq or DNase | Flags regions of open chromatin | Promoter / CRE | ENCODE; ROADMAP epigenomics |  |
| Promoter capture Hi-C | Links enhancer regions to promoters of target genes | CRE | Published datasets; 3D genome browser* | Transient interactions that may not be needed for enhancer function |
| Hi-C | Define topologically associated domains (TADs) | TAD boundaries | Published datasets; 3D genome browser* |  |
| Cap analysis gene expression (CAGE) | Marks the 5' cap of mRNA | Promoter / 5'UTR | FANTOM5 | Alternative transcripts |
| CTCF ChIP-seq | Identifies regions bound by the insulator protein CTCF | TAD boundaries | ENCODE; ROADMAP epigenomics |  |
| Transcription factor ChIP-seq | Identifies regions bound by specific transcription factors | Promoter / CRE | ENCODE; ROADMAP epigenomics |  |
| H3K4Me | Histone modification found near enhancers | CRE | ENCODE; ROADMAP epigenomics |  |
| H3K4Me3 | Histone modification found near promoters | Promoter | ENCODE; ROADMAP epigenomics |  |
| H3K27Ac | Histone modification found at active regulatory elements | Promoter / CRE | ENCODE; ROADMAP epigenomics |  |
| Expression quantitative trait loci (eQTLs) | Identifies variants that are associated with changes in gene expression | Promoter / CRE | GTEx; eqtlgen.org |  |
| Experimental perturbation | Demonstrates an impact of altering/deleting all or part an element on gene expression | Promoter / CRE | Published data | Assays may not be representative of endogenous situation |
| Northern blot, RT-qPCR, RNA-sequencing and microarrays | Detection of miRNAs | miRNAs | miRBase; published datasets |  |
| Multiple approaches | Experimentally validated miRNA–target interactions | miRNA targets | miRTarBase; published datasets |  |
| Bisuphite sequencing | Detects methylated DNA and allows identification of differentially methylated regions | Promoter / upstream gene regions | ENCODE; ROADMAP epigenomics |  |
| RNA Pol II ChIP-seq | Detection of poised polymerase II | Promoter | ENCODE; ROADMAP epigenomics |  |

#### Introns and UTRs

The definition of intronic and UTR regions is transcript dependent. In general, these should be defined using well-validated clinically relevant transcripts. Even if a well-validated transcript exists for the coding regions of a gene of interest, the UTRs may not be well defined. We therefore recommend using transcripts defined by the Matched Annotation from NCBI and EMBL-EBI (MANE) project which has integrated multiple diverse datasets to accurately define these elements. Each gene has a single ‘Select’ transcript (98% of genes) and some genes have additional ‘Plus Clinical’ transcripts.

#### Promoters

In the absence of any additional data, a minimum promoter region can be defined as the 250 bps immediately upstream of the start of the 5’UTR of a MANE Select or other well supported transcript, or a peak defined by CAGE (Cap Analysis Gene Expression) in a relevant tissue or cell type. In the same or similar biological samples, the region can be extended further upstream if a continuous region is supported by either (1) histone modifications that mark active promoter elements (H3K27Ac and H3K4Me3), (2) multiple overlapping transcription factor binding sites identified by ChIP-seq, (3) poised RNA Pol II identified by ChIP-seq, and/or (4) open chromatin as defined by ATAC-seq or DNase-seq.

#### CREs

There may be multiple ‘candidate’ CREs in the region surrounding a gene of interest. As noted above, these must have a known or functionally validated link to the gene of interest for variants within them to be interpreted clinically. We therefore outline a two-step process to identify CREs. Firstly, candidate CREs can be defined as regions of open chromatin (defined by ATAC-seq or DNase-seq), marked by histone modifications that mark active enhancer elements (H3K27Ac and H3K4Me1), and/or with evidence of multiple overlapping transcription factor binding sites identified by ChIP-seq. Candidate CREs should then be filtered to only those with experimental evidence of a link to the gene of interest, for example through chromatin interaction data (from promoter capture Hi-C), functional perturbation showing an effect on gene expression, or the presence of one or more expression quantitative trait loci (eQTLs) for the gene.

It merits repeating that enhancer and promoter usage can vary across tissues and also temporally, for example throughout development^39^. It is therefore important that when defining both promoters and CREs the above datasets should be derived from a relevant tissue or cell type for the phenotype of interest and where possible, from a relevant developmental time point.

### Predicting the impact of variants identified in candidate non-coding regions

Predicting the effect of any individual variant may not be straightforward. In Table 1 we have listed many of the mechanisms through which non-coding region variants have previously been shown to cause disease. This list is not exhaustive, and new mechanisms will be identified as more variants are identified and comprehensively studied. Often, the only way to fully determine a variant’s impact will be through functional studies (see section on PS3/BS3 below). Where *in silico* tools exist to predict the effect of certain classes of non-coding region variants, we have noted examples of these in Table 1.

### General considerations

#### Variant types covered by this guidance

This guidance is intended to cover short sequence variants (SNVs and insertions and deletions (indels) <50bps in size) to mirror the original ACMG/AMP guidelines. We do not explicitly consider larger copy number (CNV) and structural variants (SVs), which are discussed in separate existing guidelines^40^. We note, however, that multiple principles of our recommendations will apply to CNVs and SVs that do not overlap protein-coding sequence. The change in disease risk associated with variants identified through genome-wide association studies are very small and outside the scope of these guidelines.

We intend these recommendations to cover all variants identified outside of protein-coding exons, including UTRs, intronic sequence, promoters, and more distal regulatory elements. We note that canonical splice site variants (altering the GT in the first two bases of the intron or AG in the last two bases) are generally considered to be loss-of-function and are well covered by existing guidelines^41^. We caution, however, that this is not always the case and we will discuss specific scenarios where exceptions may apply.

#### Terminology

Referring to variants as either ‘coding’ or ‘non-coding’ based on where they are in genomic sequence can be an unhelpful distinction. It is much more informative to instead refer to the predicted (or possible) downstream effect of the variant, which may be to alter protein sequence, and/or to change the abundance of expressed protein. For example, both coding loss-of-function (LoF) variants and regulatory variants in non-coding regions that abolish protein expression can have equivalent downstream effects. Conversely, a UTR variant that extends the coding sequence at either the N- or C-terminus could exert a pathogenic impact through changes to the protein sequence rather than changing protein levels, and hence is best described primarily by its mechanism of pathogenesis.

### Applying the ACMG/AMP guidelines to non-coding region variants

Whilst the primary consideration for the ACMG/AMP guidelines was interpretation of variants in protein-coding regions, they were intended as all-encompassing and can be applied to interpretation of variants genome wide. These recommendations are therefore designed to sit alongside this existing guidance, noting adaptations to these rules rather than replacements.

Many of the rules from Richards *et al*. can be directly applied to variants in non-coding regions, without requiring extra considerations (Figure 3; Supplementary Table 3). These include the use of frequency information (BA1, BS1, BS2, and PM2), upweighting of confirmed *de novo* variants (PM6 and PS2), and incorporation of co-segregation evidence (PP1 and BS4). Conversely, some rules are not applicable to non-coding region variants, for example those that refer specifically to missense variants and are not further adapted here (PP2, and BP1; Supplementary Table 3).

**Figure 3:**
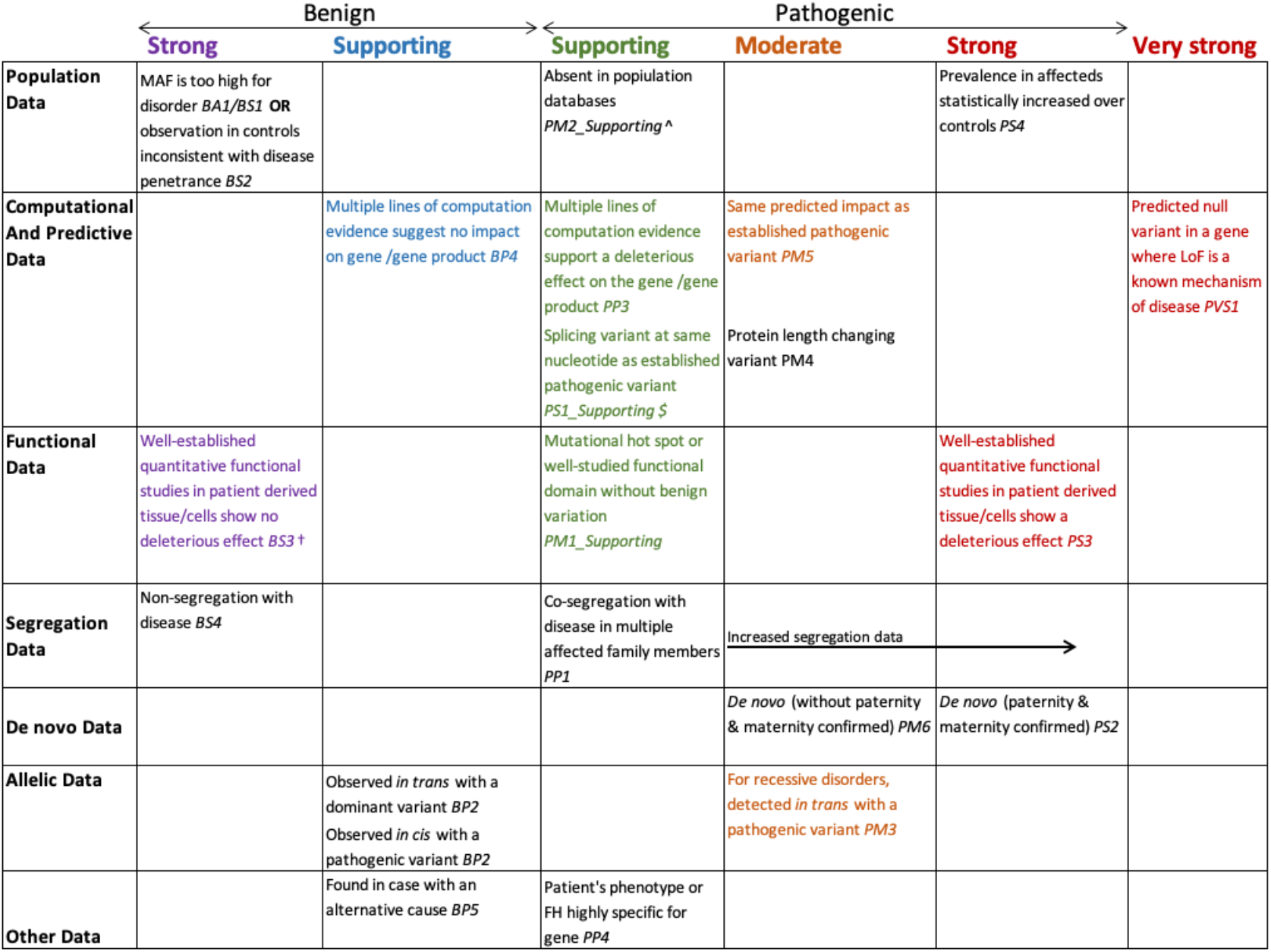
ACMG evidence framework for non-coding region variants. An adapted version of the figure from Richards *et al*.^30^ (permission granted). Rules that require no extra guidance for non-coding region variants are written in black, with those requiring extra considerations or adaptation in colour. †Should not be applied if the assay only assessed one of multiple possible mechanisms. ^Reduced to supporting following guidance from ClinGen SVI^42^. ^$^Variant must have at least as great an impact predicted by *in silico* tools.

For nine of the specific ACMG/AMP rules we recommend some modification in the way in which they are used (for example by reducing the strength to reflect lower certainty) or note extra considerations for when they are activated (Figure 3; Supplementary Table 3). Each of these are discussed in detail below.

#### PVS1

In the ACMG/AMP guidelines PVS1 is defined as “predicted null variant (nonsense, frameshift, canonical ±1 or 2 splice sites, initiation codon, single or multi-exon deletion) in a gene where loss-of-function (LoF) is a known mechanism of disease.”^30^ Given the extreme caution that is required when applying this criterion given its ‘very strong’ weighting, the Clinical Genome Resource (ClinGen) subsequently released further guidance on its application, including recommendations to decrease the strength applied to this rule under situations where the confidence in a variant being true or complete LoF is reduced^41^. Neither the original ACMG/AMP guidelines nor the updated guidance refer to non-coding region variants other than canonical splice site variants, or splicing defects that would delete one or more exons, as most variation in non-coding regions cannot be confidently predicted to lead to a null effect in the absence of experimental data. Given this, we do not recommend the use of PVS1 for variant types not covered by existing guidance. Additionally, PVS1 should not be used in combination with *in silico* prediction tools (rule PP3), as specified for canonical splice variants in previous guidance^41^.

Although PVS1 is often applied to canonical splice site variants, we caution that these do not always have a null effect; factors such as alternative splicing may mediate the pathogenic impact of these variants despite the variant causing changes to the spliced transcript^43^. Moreover, variants which disrupt splicing can have multiple impacts^44^ and/or only partial effects (see guidance below), with some aberrantly spliced transcripts resulting in a loss-of-function and others showing no discernible change or creating a functional transcript. In such situations, these consequences would receive different interpretations, and the relative dosage of each transcript is an important consideration.

Functional evidence to demonstrate a null effect of a variant in a relevant tissue is very important for non-coding region variants. In principle, these data could be used to inform either PVS1 or PS3. Under no circumstances should the same data be used for both PVS1 and PS3 as this would constitute ‘double counting’ of evidence.

#### PM1

Rule PM1, “Located in a mutational hot spot and/or critical and well-established functional domain (e.g. active site of an enzyme) without benign variation” was designed initially to capture variants within important protein domains that are critical to function. It is important to note that sequence constraint and variant effect in non-coding regions has been shown to likely be base-specific rather than consistent across larger regions^13,45^. It is therefore not appropriate to activate PM1 for a variant within a region (e.g. a UTR, or cis-regulatory element) just because multiple previous variants within that region have been shown to be pathogenic. There are, however, occasions where activating PM1 may be appropriate; for example, when a variant disrupts the binding motif of a transcription factor, perturbation of which has repeatedly been shown to be pathogenic, or where multiple known pathogenic variants are clustered within the same well-defined enhancer region, such as the ZPA regulatory sequence (ZRS) that controls expression of *SHH*^*46*^. In these instances, however, we would recommend always lowering the strength of PM1 to supporting.

In the given example of disrupting a known transcription factor binding site, if this is predicted using an *in silico* tool then this should be used to inform PP3 and should not also be used for PM1.

#### PS1

In the ACMG/AMP guidelines, PS1 is used specifically for missense variants, when a different nucleotide change results in the same amino acid change as an established pathogenic variant. Subsequent guidelines from the UK ACGS stated that PS1 could also be used “at a supporting level for splicing variants where a different nucleotide substitution has been classified as (likely) pathogenic and the variant being assessed is predicted by *in silico* tools to have a similar or greater deleterious impact on the mRNA/protein function”^47^. Whilst we support this use of PS1, we also caution that different base changes can have different effects on activation of alternative splice-sites and hence could have different impacts.

There are other specific occasions where activation of PS1 may also be appropriate, for example, uORF stop-lost variants where disruption of the same stop-codon has previously been shown to be pathogenic.

#### PM5

Similarly to PS1, PM5 is also described in the ACMG/AMP guidelines as specific to missense variants, although PM5 refers to variants disrupting the same amino acid residue, but leading to a different alternate residue. Here, we recommend using this evidence code to capture non-coding region variants that are predicted to have exactly the same impact, on the same gene, as established pathogenic variants, but themselves may not have been described before. Examples of this include upstream start codon creating variants that result in out-of-frame overlapping open reading frames (see NF1 example curation below), and near completely overlapping deletions of the same transcription factor binding site or promoter region.

We further caution on our use of the phrase ‘predicted to have exactly the same impact’. The gene specificity of regulatory elements means that identifying a variant with exactly the same impact is often not possible. For example, a variant that creates an upstream start codon in a 5’UTR may need to be created into the same context and in the same frame with respect to the coding sequence to have an identical effect, and even then, differing distances to the coding sequence may have an impact. Similarly, two variants could disrupt binding of one transcription factor, but result in opposite effects on the target gene e.g., if one variant creates a novel binding site for a paralogous transcription factor.

In general, PS1 should be used where the same base, or residue, as the previously pathogenic change is impacted, and PM5 should be used for other variants with the same predicted effect, but that are not at the same specific base/residue. If the effect of the variant is predicted using an *in silico* tool (e.g. splice region variants), then this information should inform PP3 and not PM5.

#### PM3

The PM3 rule can be used for recessive disorders when a variant is detected in *trans* with a known pathogenic variant. Non-coding region variants, in particular deeply intronic variants that impact splicing, have been identified in *trans* with coding variants^44^. Given the increased search area for possible *in trans* variants when including non-coding regions (particularly intronic regions), we sought to determine the frequency at which we would expect to observe one or more rare variants *in trans* using rare disease trios from the GEL 100,000 Genomics Project dataset.

We identified 2,016 undiagnosed trio probands with 2,714 single, rare (AF<0.5%) heterozygous pLoF variants in 794 genes in DDG2P annotated as biallelic (i.e. recessive) with a LoF mechanism. For each sample-pLoF pair, we searched for rare variants in non-coding regions of the same gene that were inherited from the alternative parent to the pLoF variant. These non-coding regions comprised intronic, 5’UTR and 3’UTR regions, and a core promoter region (200bps immediately upstream). 1,027 sample-pLoF pairs (37.8%) had at least one regulatory variant *in trans*, with a mean of 0.89 (range 0-22) identified per sample-pLoF pair (Supplementary Figure 1). As expected, the vast majority (93.9%) of *in trans* variants mapped to intronic regions, however, only seven of these passed a permissive SpliceAI threshold of 0.2. If we filter the intronic variants using this SpliceAI threshold, only 2.0% of sample-pLoF pairs had a candidate variant *in trans*.

Given the low numbers of *in trans* variants found in this analysis, we believe it is appropriate to apply PM3 as per existing guidelines for coding variants^48^. However, it is especially important to use strict allele frequency cut-offs to limit consideration to only suitably rare variants^34^ and only consider variants impacting genes that are a credible cause of an individual’s phenotype. Of note, we identified 22 variants *in trans* with one pLoF variant in the *WWOX* gene, which is an extremely large gene spanning >1.1Mbs. We also identified 16 *in trans* variants in an individual self-reported as ‘Black or Black British: African’. These examples highlight extra care required when considering genes with particularly large intronic regions or where reference datasets do not adequately match an individual’s genetic ancestry. In these instances, it may be appropriate to lower the strength of PM3. One possible approach is to calculate the likelihood of observing a similar variant (e.g. with an equivalent or greater *in silico* score) in the gene of interest, given the distribution of all scores across the gene. This approach would adjust for both region size and localised mutability. It should also be noted that non-coding region variants and hypomorphic coding variants that appear to be tolerated as homozygotes can be pathogenic when found in *trans* with a null effect coding variant^49^.

#### PS3/BS3

Functional evidence is extremely important to support either a pathogenic or a benign role for non-coding region variants. Bespoke assays are often required depending on the variant context and its predicted effect (e.g. on splicing, transcription, translation, or chromatin looping etc.). Functional assays should be designed and assessed following existing guidance from the ClinGen SVI group^50^. Below we discuss in more detail considerations for different categories of functional assays commonly used for non-coding region variants.

##### Detecting aberrant splicing events

RNAseq and/or targeted approaches enable the assessment of a number of characteristics which may be indicative of the functional impact that a variant has on normal gene expression. These include the characterisation and quantification of aberrant transcript isoforms, differential gene expression, and allelic expression imbalances^51^. Some genomic variants will cause binary changes in the measured characteristics, for example, the abolition of canonical splice sites, whereas others will cause changes in the relative ratio of normal:aberrant gene expression profiles (see guidance below on partial effects). The discovery of aberrant gene expression through RNAseq requires comparison to a control cohort (e.g. GTEx^35^) and usually also to individuals from the same sequencing and analysis process. We recommend that software used to detect aberrant splicing events has been benchmarked specifically for their discovery in the context of rare disease^52^. The technical appropriateness of biosamples for the discovery of aberrant events in the gene of interest should be considered (i.e is the gene normally expressed in this tissue?), including when using bespoke control sets. Strong weighting of PS3 for identified aberrant splicing should only be used when (1) expression profiles in the biosample used match those of the primary disease tissue of the candidate genes^53^, and (2) the sequencing data generated is appropriate for the detection of aberrant splicing events (e.g. exceeds the 95% confidence interval recommendations from MRSD (Minimum Required Sequencing Depth) using appropriate parameters for the laboratory and bioinformatics approaches applied^54^). In addition, we recommend that BS3 is only used in the absence of aberrant splicing events when there is evidence that both alleles are being expressed (e.g. data supporting heterozygous alt/ref alleles present in roughly equal quantities) and appropriate sequencing coverage has been achieved^53,54^. Even in such scenarios, caution must be taken when using BS3 due to the known cell-specific impacts of some splicing variants^55^, and, in some cases, complex alternative splicing dynamics^56^; we recommend that BS3 is not used in these situations, unless an appropriate system for functional assessment has been used.

##### MAVE approaches

multiplexed assays of variant effects (MAVEs) that classify variants as functionally normal or functionally abnormal have great potential to aid the interpretation of both protein-coding and non-coding region variants. Whilst the majority of studies to date have focused on protein-coding regions, smaller studies have profiled portions of UTRs^57^, and others have used deletion tiling to identify and study enhancers^58,59^, offering insights into the regulatory code. In general, use of MAVE data in variant interpretation should follow existing guidance^60^. Extra care should be taken when interpreting MAVE results for non-coding region variants, however, for the following reasons: (1) assays that only test a short section of a regulatory element for function do not account for regulation mechanisms that rely on neighbouring DNA, such as the formation of secondary structure, binding of co-factors, or presence of internal ribosome entry sites (IRES); (2) if only a single output is assessed, this may not be relevant for the mechanism of the variant of interest (i.e. when an RNA-seq read-out is used, but the variant is predicted to impact translation); (3) experiments may be performed in a cell type or model system where the applicability to the disease of interest is unclear. One clear limitation to current use of MAVEs is the focus of each experiment on a single gene, or even only a single exon within a gene. Collaboration through initiatives such as the Atlas of Variant Effects (AVE; www.varianteffect.org) are essential to achieving comprehensive coverage across both genes and regulatory regions.

##### Chromatin interaction assays

Chromosome conformation capture (3C) approaches can be used to identify regions of the genome that are co-restricted following chemical cross-linkage, including CREs and the promoters of their target genes. As we advise above, this information should be used to inform which variants are interpreted using these guidelines rather than being used as functional evidence to inform PS3. An exception to this is where chromatin interaction is shown to be disrupted in individuals with a variant in a candidate enhancer region (which passes the above inclusion criteria) when compared to appropriate controls, which could be used to activate PS3.

##### Reporter gene assays, e.g. luciferase assays

can be used to assess the impact of variants in promoter and regulatory regions of genes. Using the bioluminescent properties of a gene inserted into a plasmid along with a candidate regulatory region, e.g. downstream of a promoter region, one can enable assessment of the relative quantitative impact of variants in the candidate regulatory region on levels of protein production^15^. Significant disruption between reporter assays containing variant and wild-type regulatory sequences can be used to activate PS3, although with the very important caveats that this is an artificial assay system and must be appropriately validated^50^.

Many non-coding region variants may only have a *partial effect*; for example, splicing variants that affect a sub-set of transcripts, or 5’UTR variants that only partially reduce downstream coding sequence translation. For splicing variants, assays can be quantitative as described above, however, for many other assays, quantifying the precise level of an effect is difficult. Even when an effect can be quantified, whether a variant with a partial effect can cause disease is very gene dependent; for some genes only a partial reduction in functional protein can be severely detrimental, but others will tolerate partial dosage changes. Benchmarking assays across the full range of effects will therefore be important to determine gene-specific thresholds for activation of PS3.

When considering BS3, if a functional assay has not shown an effect on the gene product or its expression, care should be taken when the assay was not performed in a relevant tissue or cell type as regulation and transcript usage can be very tissue specific. In addition, BS3 should not be applied if an assay only assesses a single output but there are multiple possible underlying mechanisms (e.g. for 5’UTR variants).

Regulatory variants can act to either increase or decrease expression of a target gene. On some occasions, a functionally tested variant may cause a gain-of-function when the known mechanism for the gene is loss-of-function. BS3 could be applied when this direction of effect is inconsistent with the known gene mechanism (although we note that many dosage-sensitive genes may be both haploinsufficient and triplo-sensitive^61^).

#### PP3/BP4

The majority of widely used computational tools that predict variant deleteriousness were designed to interpret the impact of coding missense variants. These tools cannot be applied to variants in non-coding regions. There are, however, multiple tools that predict the likely impact of variants on splicing, and a few that make predictions along the complete length of a pre-mRNA transcript^62^. Comparisons between available splicing tools have been published recently^44^, showing that some tools (e.g. SpliceAI) perform well to prioritise variants with functional evidence of aberrant splicing. These tools are not further reviewed here, although we also note recent papers benchmarking the performance of CADD-splice^63^ and SQUIRLS^64^. Additionally, we caution that many tools are trained on existing canonical splice junctions, or known pathogenic/benign variants that are enriched near exon-intron junctions and hence may perform less well for deep-intronic splice variants.

*In silico* tools that can be used to predict the deleteriousness of other categories of non-coding region variants have also been recently reviewed (see Table 3 in Rajano *et al*.^65^). In addition to those mentioned, we note recent tools designed specifically for rare disease: NCBoost^66^, ReMM (Genomiser)^67^, and GREEN-DB^68^.

For genome-wide machine-learning tools that rely on a set of true positive pathogenic variants for training we caution that accurate datasets for this purpose covering non-coding regions are currently very limited. These data are biased towards certain subsets of variants, including those very close to the coding sequence and only within a small number of genes (i.e. those causing single-gene disorders). Indeed, a recent paper describing NCBoost demonstrated this regional bias^66^. How well these tools predict the pathogenicity of the full range of non-coding region variants is currently unknown. We therefore recommend extreme caution against over-interpreting the output of any genome-wide predictor.

Whilst a limited selection of *in silico* scores for non-coding region variants are accessible through widely-used annotation tools (e.g. Ensembl VEP), the vast majority must be queried individually and many are only currently available as large file downloads of pre-computed scores or through running software/scripts (Supplementary Table 4). This presents a barrier to the use of many of these tools. Of note, the GREEN-VARAN^68^ tool returns scores from a group of seven *in silico* algorithms, although it is not currently available as a web-tool.

### Example variant curations

#### NF1:c.-160C>A hypothetically identified variant in an individual with neurofibromatosis type 1

The *NF1*:c.-160C>A (ENST00000356175.7; chr17:31095159:C>A GRCh38) variant creates an upstream start codon (uAUG) in the 5’UTR of the NF1 gene. It has not been reported in ClinVar or the literature. uAUG-creating variants in *NF1* have previously been shown to cause neurofibromatosis^13,69^. This variant has the same predicted impact as these previously identified pathogenic variants: it is created into a strong Kozak consensus, and translation from this uAUG would create an upstream open reading frame (uORF) that overlaps the coding sequence out-of-frame with the canonical start site. We would therefore activate PM5. This variant is also absent from gnomAD (PM2_Supporting) and it would be appropriate to activate PP4 if the variant was identified in an individual with classic neurofibromatosis type 1 (NF1) features given the specificity of the NF1 phenotype for the *NF1* gene. If this variant occurred either *de novo* (PM6/PS2), or if there was evidence of segregation with disease at a moderate level (PP1_Moderate), it would reach a classification of Likely Pathogenic. Similarly, a functional assay demonstrating a reduction of translation in a validated cell line model, or a reduction in protein levels in an appropriate tissue sample would enable activation of PS3 resulting in a Likely Pathogenic or Pathogenic classification.

#### CFTR c.3874-4522 A>G identified in a patient with cystic fibrosis

The proband received a late diagnosis of cystic fibrosis (16-20 years old). Previous genetic testing uncovered the common c.1521_1523delCTT (p.Phe508del; chr7:117648320:A>G GRCh38) pathogenic variant in *CFTR* in a heterozygous state and was proven *in-trans* to c.3874-4522A>G (PM3). c.3874-4522A>G is absent from gnomAD (PM2_Supporting) and at the time of analysis has previously been reported in a single case of cystic fibrosis with no additional functional evidence. There is a strong phenotype-genotype correlation (PP4). MaxEntScan supported the activation of a cryptic splicing site but a number of *in-silico* splicing tools did not support that the variant would impact splicing (SpliceAI, TraP), therefore neither PP3 nor BP4 were applied. Functional assessment of splicing impact was performed using RNA extracted from whole-cell blood for the proband, showing abnormal splicing of *CFTR* with the introduction of a 125bp cryptic exon containing a stop codon. PS3_Strong was therefore applied as we were using an established method for splicing variant investigation, with appropriate controls. As we are using PS3, we would have had to exclude PP3 in our final classification had this code been used. The variant was classified as Likely Pathogenic (PS3_Strong, PM3, PM2_Supporting, PP4).

##### Emergence of new evidence

After initial classification, additional evidence became available from the literature. This included functional evidence from minigenes and patient samples^70^. As the assays performed in the initial classification were appropriate for use of PS3_Strong, there was no alteration due to this evidence. Additional families with cystic fibrosis from multiple ethnicities have also been reported to carry the c.3874-4522A>G variant, and in at least 4 of these families, symptomatic individuals are proven to carry c.3874-4522A>G *in-trans* to a proven pathogenic allele^71^. This evidence allows us to upgrade PM3 to VeryStrong, and our final classification to Pathogenic (PS3_Strong, PM3_VeryStrong, PM2_Supporting, PP4).

#### PAX6 distal enhancer variant in a patient with aniridia

The chr11:31664397 C>A (GRCh38) variant disrupts a CRE downstream of PAX6 and intronic in ELP4. The region is highly conserved and the element is identified as a ‘distal-enhancer’ by the ENCODE regulatory build (visualised on the UCSC genome browser^72^). Functional experiments show that deletion of the element disrupts maintenance of PAX6 expression^73^. Multiple in silico scores support a deleterious role (CADD=17.4; ReMM=0.985; FATHMM_MKL=0.993; PP3), and the variant is absent from gnomAD (PM2_Supporting). In silico modelling suggests the variant disrupts a PAX6 binding site which was validated through disruption of reporter expression in the lens (PS3_Moderate). The variant was also identified de novo via trio analysis (PS2) in a patient with a highly specific phenotype (PP4). Taken together, these data result in a Likely Pathogenic classification (PS2, PS3_Moderate, PM2_Supporting, PP3, PP4).

## DISCUSSION AND FURTHER CONSIDERATIONS

Here, we have outlined considerations for adaptation of the ACMG/AMP guidelines for variant interpretation to variants identified outside of protein-coding regions of the genome. These recommendations have been carefully reviewed and refined by an expert panel and extensively tested by clinical variant scientists.

It is clear that our knowledge of the impact of non-coding region variants in rare disease is a fast-evolving field, which makes it complex to provide comprehensive guidance that will fit every possible scenario. We have therefore tried to provide general guidance that can be applied to most variant types, and have included specific examples of how to apply this guidance in practice. We hope that these recommendations will enable increased interpretation of non-coding region variants and catalyse the discovery of additional examples of disease-causing variant types, which will in turn inform further revisions to this guidance. To enable this continued learning and refinement of these guidelines, we encourage the sharing of variant data, including for variants that are not initially classified as pathogenic, for example by submitting classified variants to ClinVar^31^ and enabling access to individual-level data and through DECIPHER^72^.

During testing of these guidelines, it became clear that one of the largest barriers to widespread adoption is access to the epigenetic data and *in silico* scores required to interpret non-coding region variants. We are in desperate need of accessible tools that allow better visualisation of these data by individuals not well-versed in bioinformatics, ideally allowing for queries by cell type/tissue, to allow transparent curation and reproducible interpretation of these data. We also need more research aimed at deciphering the full ‘regulatory code’ and development of *in silico* prediction tools for non-coding variants not trained on limited pools of known pathogenic variants. It was also clear from feedback that there is substantial appetite for educational webinars and workshops around various aspects of using the guidelines (Supplementary Table 2). This includes training on finding and evaluating functional data which may use assays that are unfamiliar. We are actively engaged in developing these in the hope they will increase usability and adoption.

A substantial barrier to our understanding of the role of non-coding region variants in rare disease to date has been the lack of statistical power when searching for enrichment of variants across and between different regulatory regions and/or variant classes. This is due to multiple factors, including (1) the majority of non-coding region variants having little or no effect, (2) a lack of clarity on how to subdivide the genome for genome-wide scans, (3) potential opposing actions of different variants, and (4) a lack of large-scale whole genome sequenced datasets derived from different genetic ancestry groups with linked phenotypic data. Conversely, much of our success to date in identifying disease-causing non-coding region variants has been using phenotypically highly-selected cohorts, where only one or a very small number of genes are suspected as being involved. It is likely that this approach will continue to be successful at identifying new disease-causing variants in non-coding regions.

The full range of mechanisms through which variants in non-coding regions cause, and contribute to the risk of genetic disease remains unknown. It is likely, however, that many regulatory variants have smaller effects than those impacting protein sequence, and that these effects may be highly tissue-specific. Whilst for some extremely dosage sensitive genes even partial effect variants will cause severe disease (as has been shown for MEF2C^15^), in others, single variants with only a moderate effect will be insufficiently deleterious or may only cause disease in a single tissue or organ. For example, in *PRPF31* disease-causing variants causing significantly reduced expression of a single allele can be incompletely penetrant^73^. The penetrance of these variants can be modified by the relative expression of the other allele, or regulatory variants could themselves modify the penetrance of damaging protein-coding variants^74^, and/or cause disease only in combination with other variants. Further research is needed to fully elucidate the frequency and impact of these different mechanisms.

The majority of non-coding region variants would be expected to either decrease or increase the protein product of an impacted gene, whether through affecting transcription, or post-transcriptional regulation mechanisms. It is therefore expected that these variants would primarily impact genes that are dosage sensitive. There are, however, exceptions, such as uAUG-creating variants in 5’UTRs that elongate the coding sequence at the N-terminus, which could negatively impact non-dosage sensitive transcripts. Furthermore, there may be occasions where a gene does not appear to be constrained against coding loss-of-function variants (so may not be considered haploinsufficient), but where regulatory variants that decrease protein levels could be deleterious. For example, if a compensatory mechanism relies on nonsense mediated decay^75^.

In these guidelines, we have primarily discussed variants that impact existing regulatory regions, however, there are examples of disease-causing variants that act through creating novel regulatory elements. For example, a recent paper discussed a SNV that created a new promoter leading to dysregulation of genes in the human α-globin locus^76^. We have also not discussed variants in non-coding genes in detail, however note examples of identified pathogenic variants in this area of active research (Table 1). The importance of variants in both creating novel regulatory elements and impacting non-coding genes will increasingly be recognised and ensuring that these guidelines continue to be appropriate for these variant types will be an important consideration in future revisions.

It should be recognised that non-coding regions can be very large. Whilst 5’UTRs average only 197 bps, 3’UTRs are on average approximately the same size as the protein-coding sequence (1,775 bps for 3’UTRs vs 1,745 bps for protein-coding sequence). The average combined size of intronic regions, by contrast, is 34-fold larger than protein-coding sequence at 59,220 bps (region lengths calculated across all MANE Select v0.95 transcripts). Including non-coding regions in the search for likely disease-causing variants therefore dramatically increases the genomic search space. We have not here recommended decreasing the strength of evidence applied to *de novo* variants for those found in non-coding regions, but more research is required into the relative rates of *de novo* occurrence across these different regions to determine whether this should be accounted for in future revisions of these guidelines.

As our knowledge of disease-causing mechanisms, and the size of both available phenotype-linked sequencing data and MAVE datasets profiling non-coding regions expand, our ability to fully interpret variants in non-coding regions for a role in both rare and common diseases will continue to increase. Given this increasing knowledge, it may be necessary to revisit and re-interpret variants initially designated as ‘research only’. This in turn will, however, catalyse the return of a definitive genetic diagnosis to ever increasing numbers of individuals with rare diseases.

## Data Availability

All data produced in the present work are contained in the manuscript.

## ACKNOWLEDGEMENTS

We would like to thank the Clinical Genome Resource (ClinGen) Sequence Variant Interpretation (SVI) working group for their useful input and guidance. NW is supported by a Sir Henry Dale Fellowship jointly funded by the Wellcome Trust and the Royal Society (Grant Number 220134/Z/20/Z) and by grant funding from The Rosetrees Trust (Grant Number H5R01320). JME is supported by a postdoctoral research fellowship from the Health Education England Genomics Education Programme (HEE GEP). DB and JL are supported by DB’s NIHR Research Professorship (RP-2016-07-011). WGN is supported by the Manchester NIHR Biomedical Research Centre (IS-BRC-1215-20007) and JCT is supported by the Oxford NIHR Biomedical Research Centre. DRF is supported by the MRC University Unit award to the University of Edinburgh for the MRC Human Genetics Unit and by the Wellcome Strategic Award “Transforming Genomic Medicine Initiative” (200990/Z/16/Z). JI is supported by a National Health and Medical Research Council Career Development Fellowship (#1162929). AODL is supported by the National Human Genome Research Institute grant U01HG011755. The ClinGen SVI as well as SMH and HLR are supported by the National Human Genome Research Institute grant U24HG006834.

This research was made possible through access to the data and findings generated by the 100,000 Genomes Project. The 100,000 Genomes Project is managed by Genomics England Limited (a wholly owned company of the Department of Health and Social Care). The 100,000 Genomes Project is funded by the National Institute for Health Research and NHS England. The Wellcome Trust, Cancer Research UK and the Medical Research Council have also funded research infrastructure. The 100,000 Genomes Project uses data provided by patients and collected by the National Health Service as part of their care and support.

The views expressed are those of the authors and not necessarily those of the NHS, the NIHR or the Department of Health.

## AUTHOR CONTRIBUTIONS

NW, JME, and SMH conceived the project and wrote the initial draft recommendations. JWA, DB, SE, DRF, WGN, and JCT formed the expert review panel. JL and HCM analysed and interpreted data. RDB, SB, CC, KD, SE, CDF, JI, NK, AODL, SCR, HLR, ER, MSB, MW, JCW, and CFW tested the guidelines and provided feedback.

---

**Supplementary Figure 1:**
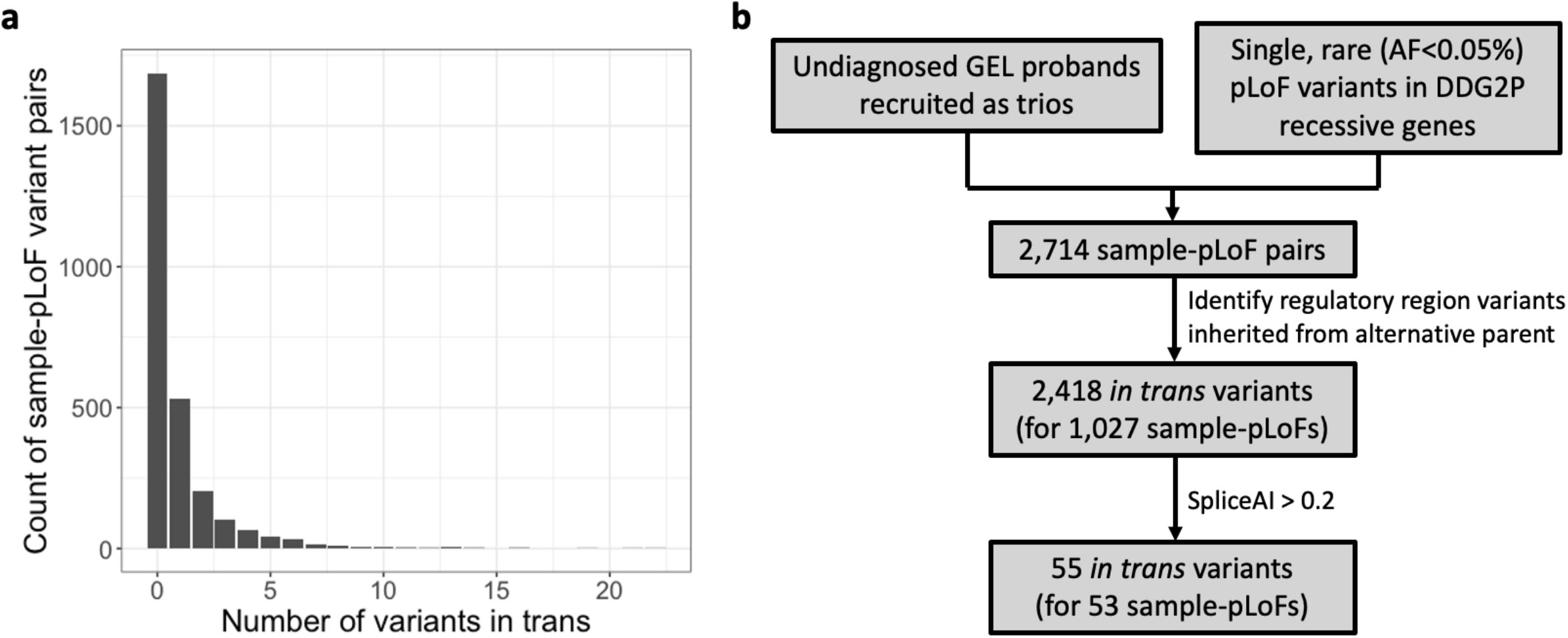
Identifying regulatory variants *in trans* with pLoF variants in GEL. (a) The number of variants identified in trans per sample-pLoF pair. One or more *in trans* variants were found for 37.8% of pLoF variants. (b) A flow diagram of the approach.

